# Pre-Service Education and Continuous Professional Development on Female Genital Mutilation/Cutting for maternal health professionals working in OECD countries: A Scoping Review Protocol

**DOI:** 10.1101/2022.08.16.22278598

**Authors:** Lisa Apini-Welcland, Marina A.S. Daniele, Lucia Rocca-Ihenacho, Christine McCourt

## Abstract

The aim of this scoping review is to map the available evidence on pre-service education and continuous professional development (CPD) for maternal health professionals providing services to pregnant women with Female Genital Mutilation/Cutting (FGM/C) in OECD countries.

FGM/C is a form of gender-based violence and has become a global phenomenon due to changing patterns in migration flows. Pre-service education curricula and CPD for maternal health professionals need to ensure FGM/C inclusion in order to provide quality services in high-prevalence countries or those serving as home for diaspora communities.

Inclusion criteria are studies, training curricula for the education of midwives, doctors or other health professionals providing maternity services, protocols and guidelines on FGM/C training, other online FGM/C training resources from OECD countries. Documents from 2010 onwards will be included. Studies in English, Spanish, French, Italian, Portuguese and German are eligible for inclusion.

The search will be carried out using keywords derived from the Population-Concept-Context (PCC) framework and entered into three databases. A grey literature search will be conducted to identify additional material, policy documents, training resources and PhD theses. The search strategy will be supplemented by focused searching for guidelines and other online resources on FGM/C and CPD activities as well as study curricula. Key personnel from education institutions, professional associations, regulatory bodies and FGM/C experts will be contacted to contribute to the review based on their knowledge and experience. All eligible material will be objectively summarized and transferred into a standardized data extraction form.

Findings will be collated by type, search studies or educational curricula to draw overall conclusions about the status of professional training opportunities in OECD countries.

## Introduction

Worldwide more than 200 million women and girls have experienced Female Genital Mutilation/Cutting (FGM/C) and additional estimates suggest that over three million girls annually are at risk of this human rights violation (WHO 2022). FGM/C constitutes a form of violence against women and gender-based violence (UN Women 2017). Power imbalances and deeply rooted cultural and social structures, values and norms, far too often perpetuated by cultural denial and silence, are underlying factors of this expression of gender-based violence (Pandea et al. 2019). From a global perspective, the picture often presented is incomplete and often leads to stereotyping certain Sub-Saharan African countries. Data from large-scale representative surveys or other sources of national representative data reports that FGM/C is practiced in 33 African communities (End FGM European Network 2020 et al, UNFPA 2021). It is important to acknowledge that there are also certain ethnic groups in Latin America, Eastern Europe, the Middle East and in Asian countries practicing FGM/C (Ibid). Furthermore, changing patterns in migration flows to North America, Europe, the Gulf and Australia complement the bigger picture with a growing body of evidence that FGM/C is equally practiced in diaspora communities, leading to the conclusion that FGM/C has become a global phenomenon (Flahaux & DeHaas 2016, End FGM European Network et al 2020).

### Data collection on FGM/C

While FGM/C is known to be performed in high-income countries (HICs), there is still a lack of quality and reliable data on the practice (Turkmani et al 2020). As the majority of high-income countries are culturally diverse, comprehensive data on ethnicity and health is needed to inform policy development and guide service development in order to target risk factors or specific health conditions among certain ethnic groups (Bignault & Hagshenas 2005). On the other hand, every human being has the right to equity and should be protected against discrimination. That’s why the Council of Europe, Europe’s leading human rights organization, outlined with the Race Equality Directive 2000/43/EC the guiding principles to equal treatment for all people, irrespective of race and ethnic origin (President of the Council of Europe 2000). So far, there are only three countries in Europe (Finland, Ireland, UK), where public bodies are obliged to collect equality data (Hannigan et al 2019). National prevalence data is one essential component to measure the extent of FGM/C at country level. In addition, medical records, e.g. patient records or maternity registers, play a vital role in enabling us to visualize the existence of FGM/C within a healthcare system. Some countries (e.g. Belgium, France, Ireland, Netherlands, Portugal, Sweden and UK) have already started to collect information about FGM/C in medical records, but as tools are still new, future evaluations are required and limitations (e.g. under-recording due to lack of knowledge) need to be addressed (EIGE 2013).

### Quality of Education and Service Provision

There is a rise in studies assessing healthcare professionals’ knowledge, attitudes and practices regarding the care for women with FGM/C in high-income countries as services are increasingly getting challenged to provide appropriate care pathways. The process of diagnosing and managing FGM/C is often accompanied by challenges, as a result of knowledge gaps or lack of corresponding skills. A Systematic Review by Balfour et al (2016) on interventions for healthcare providers to improve treatment and prevention of FGM/C highlighted that training effects, obstetric outcomes and user satisfaction are rarely evaluated. In addition, FGM/C expert Richard (2017) reinforced the importance of integrating FGM/C into health professionals’ training curricula. This is a long outstanding recommendation, although progress is difficult to track. The World Health Organization (WHO) has produced practical guidance on how to integrate FGM/C into nursing and midwifery curricula, which is reportedly based on a preliminary review of existing curricula. The authors state that they identified training gaps, as well as an increasing demand for a systematic approach. However, they do not report on the process and findings of this review (WHO 2022). Additionally, prior to undertaking this project I conducted a preliminary search and did not identify any systematic reviews or scoping reviews on Medline and the Cochrane Database of Systematic Reviews. This review seeks to map the available evidence on pre-service and continuing professional education activities for maternal health professionals providing services to pregnant women with FGM/C in OECD countries. Findings from this work will inform an exploratory case study on maternity service provision for women with FGM/C in a University Hospital in Germany and connect data from a situation analysis with training needs as part of a PhD thesis.

### The Emphasis on OECD countries

The Organization for Economic Cooperation and Development (OECD) has the ultimate goal to shape policies that promote equality, prosperity, opportunity and well-being for all of its 38 member states (OECD 2022). In 2019 the OECD Health Policy Studies “Health for Everyone?” analysed inequalities in health and health systems, which encouraged an important dialogue of how societies could become more inclusive (OECD 2019). It is crucial for OECD members to understand that unmet needs for care are systematically more prevalent among lower income groups (lbid). That’s why members and partners of the OECD need to collaborate closely on global issues in order to promote and initiate reforms with the underlying foundation of shared values and wisdom (OECD 2022). Unfortunately, inequalities in health related to migration or ethnicity are not yet considered in the report but reinforce the need that appropriate maternity care pathways and quality services for women with FGM/C are needed as it is essential to integrate FGM/C into health professionals training curricula and education activities.

## Review question

What Pre-Service and Continuous Professional Education activities on FGM/C are available for maternal health professionals in OECD countries?

Additionally, the review will explore the following sub-questions:

i. How many hours of training do maternity healthcare students or professionals receive on FGM/C?
ii. What are the training formats and contents covered?
iii. Which competencies can be achieved? How are they measured and evaluated?
iv. What is the effect of training on the clinical competence, obstetric outcomes and user experience?

## Eligibility criteria

### Participants

The focus of this review is on maternal health professionals as a group of skilled health workers, who are educated, trained and regulated to national and/or international standards, competent to provide maternal and newborn health services (WHO 2018). Maternal health professionals are part of interdisciplinary teams with the aim to promote and support physiological processes as well as to identify and respond to medical complications and provide emergency procedures when needed. Beside these specific competencies across the continuum of care for pregnant women and newborn infants, general competencies such as professional accountability and the provision and promotion of evidence-based, human-rights based and high-quality services are key pillars of maternal health professionals’ codes of conduct (ICM 2018, WHO 2018). This review is focused on the following maternal health professionals, in alphabetical order: anaesthetists, general practitioners, gynaecologists, midwives, nurses, obstetricians and paediatricians.

### Concept

The concept of this review explores pre-service education curricula and continuous professional development education materials for maternal health professionals. It is an essential prerequisite that education for maternal health personnel takes account of population needs and health service demands (Evans et al 2016). Pre-service education and CPD activities need to balance the pressure of providing curricula that meet the standards of international labour markets, as well as responding to local needs (WHO 2016). Competency-based learning and interdisciplinary learning should equip maternal health workers to effectively take account of social determinants of health in relation to the care of women with FGM/C. Interdisciplinary collaboration and sustainable approaches are urgently needed. This review aims to explore training curricula and CPD materials on FGM/C for maternal health students and professionals provided by the public, private not-for-profit and private for-profit organisations and institutions. The training format may include lectures, skills lab sessions, workshops, webinars or e-learning activities at undergraduate or postgraduate level. It will not include materials such as textbooks or information articles which might have the potential to be used as resources within or by training programs, but which are not explicitly designed and presented as educational materials or curriculum components.

### Context

The context of this review covers all member states of the Organisation of Economic Cooperation and Development, as the ultimate goal of this alliance is to shape policies for the equality and well-being of every person, including every woman with FGM/C living in one of these countries. As it is crucial to leave behind persistent stereotypes which exclusively associate FGM/C with African countries, this research recognizes FGM/C as a global phenomenon and explores its occurrence in high-income economies with a high human development index, organized within the OECD system.

### Types of Sources

This scoping review will consider qualitative, quantitative and mixed-methods studies as well as any other study or report investigating education activities on FGM/C in maternity care in one or more of the 38 OECD countries. Pre-service education curricula and CPD trainings for maternal health professionals, not specified to a particular training mode, will also be included. Documents may vary between different sources as they might have been originated by public, private or the NGO sector. Duplicate articles, opinion papers, news articles and articles without full text available will be excluded from the review. The search strategy is restricted to evidence from 2010 onwards in order to ensure learning lessons from the early stages of FGM/C provision have been incorporated and ensure a certain level of currency. Language proficiencies covered by the research team are English, Spanish, French, Italian, Portuguese and German. In case possibly eligible material is identified in another language, we will approach key informants from academia, clinical settings or professional associations for support.

## Methods

The proposed scoping review will be conducted in accordance with the PRIMSA Extension for Scoping Reviews (Tricco et al 2018). Scoping reviews are specifically designed to systematically identify and map the breadth and depth of available evidence, summarize and disseminate findings. Using this method, relevant areas in need of further investigation should be determined (Arksey & O’Malley 2005, Munn et al 2022). This approach to evidence synthesis was chosen in order to explore the concept of Pre-Service Education and Continuous Professional Development activities on FGM/C in maternity service provision. We also aim to identify key characteristics of the concept and examine provision across the context of OECD countries.

### Search strategy

The search strategy will aim to locate both published and unpublished sources. An initial database search via Google Scholar and Medline was undertaken using keywords such as female genital mutilation, female genital cutting, healthcare professionals, education and continuous professional development to get an overview of the topic. Further keywords contained in the titles and abstracts of relevant articles and index terms were used to develop a full search strategy for CINAHL, Embase and Medline (see Apendix I). The search strategy will be applied to all databases and/or other sources of information. Reference lists of all included records will be screened to identify further studies and documents on FGM/C pre-service education and CPD activities for maternal health professionals from OECD countries. Additionally, a grey literature search via OpenGrey and the E-Theses Online Service (EThOS) will be undertaken in order to identify policy documents, training materials and PhD theses relevant to the topic. Studies published in English are eligible for screening. Additionally, education materials or other training resources in French, German, Italian, Spanish and Portuguese will be included as the review team has respective language proficiencies. In case of language restrictions or data gaps, midwives and medical associations as well as representatives from academia or other relevant key persons will be contacted for further information or clarification. Studies published with effect from 2010 will be included to ensure learning lessons from the early stages of FGM/C service provision have been incorporated and a certain level of currency ensured.

The databases to be searched include CINAHL and Embase via EBSCOhost and Medline via PubMed. Sources of unpublished studies or grey literature to be searched include OpenGrey, the E-Theses Online Service (EThOS), websites from WHO, UNICEF and UNFPA, official OECD documents as well as from the European Union applying to an OECD country, and individual country reports from an OECD country (public, private not-for-profit and private for-profit organisations and institutions and professional associations.

### Source of Evidence selection

Following the search, all identified citations will be collated and uploaded into ProQuestRefworks citation management software (Clarivate, Michigan, USA). Subsequently, studies will be uploaded into Covidence (Veritas Health, Melbourne, Australia) and duplicates removed. Two independent reviewers will screen titles and abstracts against the inclusion criteria. Records retained based on screening will be retrieved with full text. Disagreement between the two reviewers will be resolved through discussion or by consulting another member of the review team. Reasons for the exclusion of sources of evidence after reviewing the full text will be recorded and reported. The results of the search and selection process will be reported in full in the final scoping review article, and presented through a Preferred Reporting Items for Systematic Reviews and Meta-analyses extension for scoping review (PRISMA-ScR) flow diagram (Tricco et al. 2018).

### Data Extraction

The data extraction tool was developed by the review team (see Appendix II). The phases in the development of the tool are represented in Figure 1. Firstly, each member of the review team will test the tool individually and extract data from three included studies. After that the review team will meet, discuss findings and amend the tool, where necessary. This approach aims to test the feasibility of the tool and consistency within the team. Data will then be extracted from all included sources by the main reviewer. The review team already conducted an initial pilot of the tool during the development process. Based on this, it seemed feasible to subdivide the findings into more specific areas and to include graded recommendations.

**Figure 1:**
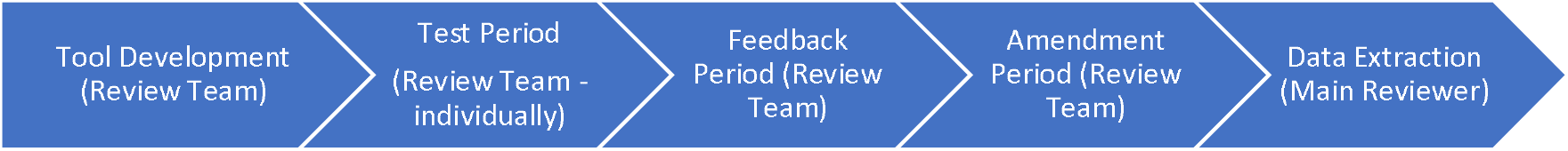
Data Extraction: Tool modification and different extraction stages

The extracted data will include specific details about the population, concept, context, study methods, if relevant, or other information sources as well as the development process (for curricula) or key findings relevant to the research questions. The draft tool contains information about the authors and study title, curriculum initiation year or training type, OECD region and country, training aim and objective, target maternal health professions, FGM/C indicator related to the availability and quality of services (UNFPA & UNICEF 2020) and findings related to the effect of the training on health professionals (WHO 2022), leadership, policy makers, educators and regulatory bodies. An additional column will allow space for recommendations. We will state whether each recommendation was derived based on the evidence provided by the study, or whether it was made by the study authors or the review team. If appropriate, authors of sources will be contacted to request missing or additional data, where required. During the selection and data extraction processes the tool may be iteratively modified and revised in order to ensure all information required to answer the research question is included and clearly presented.

## Data Analysis and Presentation

All evidence presented will directly respond to the review objectives and questions. Data extracted during the review will be processed and presented in a tabular or diagrammatic form. The tabulated and/or charted results presentation will be accompanied by a narrative summary, which will describe how the findings relate to the objectives and questions of the scoping review. An assessment of study quality is not necessary during a scoping review and has not been included in this protocol, as the overall aim is to map available evidence and identify gaps for further research.

## Supporting information

Search Strategy

Data extraction tool

Data extraction tool curricula review

## Data Availability

All data produced in the present work are contained in the manuscript.

## Acknowledgements

Dr. Jasmine Abdulcadir supported the research team with her expertise on FGM/C and maternity care, especially with a focus on high-income countries. Andreia Soares Goncalves guided the main researcher with her professional experience on scoping reviews from the perspective of a midwife and PhD student. This review contributes towards a PhD degree (LAW).

