## Supplementary material for "Pre-Service Education and Continuous Professional Development on Female Genital Mutilation/Cutting for maternal health professionals working in OECD countries: A Scoping Review Protocol": Search Strategy

### Appendices

### Appendix I: Search strategy (conducted on Embase at 7. April 2022)

| **Search** | **Query** | **Records retrieved** |
| --- | --- | --- |
| # 1 | female genital mutilation.m_titl. | 1,082 |
| # 2 | limit 1 to (abstracts and english language and yr="2010 -Current") | 586 |
| # 3 | education.m_titl. | 132,223 |
| # 4 | limit 3 to (abstracts and english language and yr="2010") | 2,837 |
| # 5 | continuous professional development.mp. [mp=title, abstract, heading word, original title, keyword heading word, floating subheading word, candidate term word] | 658 |
| # 6 | limit 5 to (abstracts and english language and yr="2010 -Current") | 492 |
| # 7 | health professional.mp. [mp=title, abstract, heading word, original title, keyword heading word, floating subheading word, candidate term word] | 12,822 |
| # 8 | limit 7 to (abstracts and english language and yr="2010 -Current") | 9,016 |
| # 9 | midwives.mp. [mp=title, abstract, heading word, original title, keyword heading word, floating subheading word, candidate term word] | 18,110 |
| # 10 | limit 9 to (abstracts and english language and yr="2010 -Current") | 10,998 |
| # 11 | 4 or 6 | 3,326 |
| # 12 | 1 and 11 | 0 |
| # 13 | 1 and 3 | 14 |
| # 14 | 1 and 5 | 0 |
| # 15 | 8 or 10 | 19,876 |
| # 16 | 1 and 15 | 41 |
| # 17 | 3 and 16 | 1 |
| # 18 | limit 13 to (abstracts and english language and yr="2010 -Current") | 9 |
| # 19 | female genital mutilation.ab. or female genital mutilation.ti. or female genital cutting.ab. or female genital cutting.ti. or female circumcision.ab. or female circumcision.ti. or FGM.ab. or FGM.ti. | 3,069 |
| # 20 | education.ab. or education.ti. or continuous professional development.ab. or continuous professional development.ti. or training.ab. or training.ti. or skills.ab. or skills.ti. | 1,300,602 |
| # 21 | limit 20 to (english language and yr="2010 -Current") | 864.460 |
| # 22 | 19 and 21 | 390 |
| # 23 | midwives.ab. or midwives.ti. or health worker.ab. or health worker.ti. or health professional.ab. or health professional.ti. or doctor.ab. or doctor.ti. | 109,372 |
| # 24 | 22 and 23 | 46 |
