## Supplementary material for "Pre-Service Education and Continuous Professional Development on Female Genital Mutilation/Cutting for maternal health professionals working in OECD countries: A Scoping Review Protocol": Data extraction tool

**Review aim:** To map the available evidence on pre-service education and continuous professional development for maternal health professionals providing services to pregnant women with FGM/C in OECD countries

**Review questions:** What kind of Pre-Service and Continuous Professional Education activities on FGM/C are available for maternal health professionals in OECD countries?

i) How many hours of training do maternal health professionals receive on FGM/C? ii)What are the training formats and contents covered? Iii)Which competencies can be achieved? How are they measured and evaluated? iv)What is the effect of training on the clinical competence, obstetric outcomes and user experience?

**Reviewer/Date:**

**Name of author(s):**

**Publication year:**

**Document type (research, curriculum, policy, other):**

**Citation details:**

**OECD-Region:** Americas	Asia	Europe	Oceania

**Study or source location:**

**Study or curriculum/policy aim:**

**Methodology or learning outcomes:**

**Study population/intended course participants:**

**Study or policy concept/education model:**

**Training mode:** modular	hybrid	integrated	others

**Training duration:**

**Training activity:** mandatory	voluntary

**FGM training indicator:** FGM prevalence

FGM drivers and types

FGM legislation and health policies

KAP among health professionals

Data on service users’ expectations

Evaluation on FGM training results

**General findings:**

**Findings related to health professionals:**

**Findings for leadership:**

**Findings for policy makers, educators, regulatory bodies:**

**Important remarks:**

**Recommendations from the Evidence:**

**Recommendations from the Author:**

**Recommendations from the Review Team:**
